# Role of Artificial Intelligence-Powered Conversational Agents (Chatbots) in Musculoskeletal Disorders: A Scoping Review Protocol

**DOI:** 10.1101/2024.08.28.24312687

**Authors:** Joaquín González Aroca, Laura Vergara-Merino, Camila Micaela Escobar Liquitay, Humberto Farías A, Jorge Olivares Arancibia, Álvaro Puelles

**Affiliations:** School of Kinesiology, Universidad de La Serena, La Serena, Chile; Department of Traumatology and Orthopedics, Pontificia Universidad Católica de Chile, Chile; Research Department, Instituto Universitario Hospital Italiano de Buenos Aires, Buenos Aires, Argentina; Department of Industrial Engineering, Universidad de La Serena, La Serena, Chile; AFySE Group, Research in Physical Activity and School Health, School of Pedagogy in Physical Education, Faculty of Education, Universidad de las Américas, Santiago, Chile

**Keywords:** Musculoskeletal Disorders, Artificial intelligence, Chatbot

## Abstract

**Introduction:** Musculoskeletal disorders (MSDs) represent a significant global health burden, leading to substantial disability and socioeconomic impact. With the rise of artificial intelligence (AI), particularly large language models-driven conversational agents (chatbots), there is potential to enhance the management of MSDs. However, the application of AI-powered chatbots in this population has not been comprehensively synthesized.

**Objective:** To explore the current and potential use of AI-powered chatbots in the management of MSDs. The review will map out the targeted diseases, the purposes of chatbot interventions, the clinical tools or frameworks utilized in training these systems, and the evaluated outcomes in clinical settings.

**Methods:** This scoping review will follow the PRISMA-ScR guidelines, with a comprehensive search across multiple databases including Medline (Ovid MEDLINE), Embase (Ovid), ISI Web of Science (wos; clarivate) and ClinicalTrials.gov. Studies involving adults with MSDs, regardless of publication status, language, or year, will be included. The scoping review will exclude studies using non-AI chatbots or human health coaches. Data extraction and synthesis will focus on demographic characteristics, chatbot methods, outcomes, and thematic analysis.

**Ethics and dissemination:** Formal ethical approval is not required as this study involves neither human participants nor unpublished secondary data. Findings of the scoping review will be disseminated through professional networks, conference presentations and publication in a scientific journal.

**Strengths and limitations of this study:**

- We developed a search strategy with high sensitivity to retrieve studies
- This scoping review will not exclude studies by publication status, language, or year of publication.

## Introduction

Musculoskeletal disorders (MSDs) are a significant global health concern, ranking as the second cause of non-fatal disability in 2020 and affecting over 1.63 billion individuals worldwide.^1^ The burden of MSDs extends beyond physical limitations, often leading to substantial disability, with MSDs topping the list in terms of years lost to disability, according to the latest Global Burden of Disease report.^2^ The enduring pain and disability associated with MSDs can also precipitate long-term psychological consequences.^3^ Additionally, the considerable healthcare costs and the strain on workforce availability further compound the societal burden of these conditions.^4^

Amidst these challenges, artificial intelligence (AI) emerges as a transformative tool, reshaping how we process information and augmenting decision-making processes through problem-solving, reasoning, and learning. AI encompasses a range of methods, including machine learning, deep learning (DL), and natural language processing (NLP). Large Language Models (LLMs) represent a specific type of AI that employs DL and extensive datasets to comprehend, summarize, generate, and forecast new text-based content ^5-7^. In the healthcare field, AI applications are diverse, ranging from machine learning algorithms that assist in diagnostic tools to LLMs that dialogue with patients. ^8^ This ability to engage in real-time dialogue with patients not only improves accessibility to healthcare services but also empowers patients by offering immediate, reliable information. Particularly, NLP models based on LLMs have gained prominence, enhancing their capabilities by generating and understanding human language through learned patterns rather than relying solely on predefined rules.^9-10^ From these models, the use of Chatbots arise These systems engage users in dialogue, –typically online– and generate responses based on analyzed inputs and accessed knowledge.^11^

The use of chatbots has been reported in various aspects of health care, such as cancer ^12^, behavioral change ^13^, and psychiatry ^14^, among others. Particularly in musculoskeletal care, chatbots have been studied in people with chronic pain ^15^, shoulder arthroplasty ^16^, and back pain ^17^. However, to our knowledge, there is no evidence synthesis about the use of chatbots in this population.

This scoping review aims to provide an overview of the current and potential use of Artificial Intelligence-Powered Conversational Agents (chatbots) in people with MSD.

## Review questions

1. In the context of MSDs, in which diseases and for what purpose have chatbots been used? (eg. medication reminders, exercise-based treatments reminders, education and motivation)
2. What clinical tools or conceptual frameworks are serving as benchmarks in training chatbots with artificial intelligence algorithms for application in clinical settings related to musculoskeletal conditions?
3. What outcomes are evaluated when implementing AI chatbots for the management of musculoskeletal disorders in clinical settings?

## Methods and analysis

This scoping review will be developed following the Preferred Reporting Items for Systematic Reviews and Meta-Analyses extension for Scoping Reviews (PRISMA-ScR). ^18^

### Eligibility Criteria

This review will include all primary studies that assess the use of AI chatbots in adults (older than 18 years) with a musculoskeletal disease in any setting. Musculoskeletal disorders refer to a diverse set of conditions affecting muscles, bones, joints, and related tissues, with symptoms that can vary in duration.

Articles will be excluded if they (1) used chatbots that did not incorporate AI (eg, chatbots and computerized coaches that were not conversational and without machine learning capabilities), (2) used human health coaches conversing with users through messaging platforms and (3) included participants that experience pain due to particular pathological causes (such as neurological disease, malignancy, inflammatory disease) will be excluded.

We will not exclude by status of publication, language, or year of publication.

Narrative reviews, letters to editors, or any non-original study will be excluded from this scoping review.

### Search Methods

We will search the following databases from inception with no restrictions on date, language or publication status:

1. Medline (Ovid MEDLINE);
2. Embase (Ovid);
3. ISI Web of Science (wos; clarivate);
4. ClinicalTrials.gov (www.clinicaltrials.gov); For detailed search strategies, see **Appendix 1**

### Selection of studies

All identified records will be uploaded to the Rayyan web application after duplicates removal. Prior to the screening process, a pilot test of the proposed eligibility criteria will be performed using three to ten articles to solve any possible disagreements regarding the selection process. Two independent reviewers will then screen titles and abstracts, and further select the studies by reading the full text of potentially eligible studies. If there is a discrepancy in any step of the selection process a third reviewer will decide whether the study meets the eligibility criteria. The search results and the reasons for exclusions will be recorded and reported in a PRISMA-ScR flowchart.

### Data extraction process

Two independent reviewers will extract data from the selected articles into a predefined template. For each study we will extract the Author, year of publication, country of origin, objectives, study population and sample size, intervention or exposure, measured outcomes, details of these outcomes, musculoskeletal disorder, and those key findings that relate to the question of this review. In case of discrepancies, a third reviewer will participate in this process.

### Collating, summarising and reporting the results

The findings will be summarized in accordance with PRISMA-ScR guidelines, focusing on descriptive and thematic analysis. Descriptive elements will cover demographic characteristics, research country, publication details, and bibliometric data. This includes musculoskeletal diagnosis, description of the AI chatbot method, purpose of the AI chatbot, nature of the dataset (eg, size, data cleaning or preparation methods), the outcomes assessed, knowledge user engagement and healthcare setting of the study. Additionally, we may conduct further bibliometric analysis, such as cocitation networks among authors and countries using VosViewer. ^19^ Furthermore, a thematic summary will be provided to emphasize the main themes identified in the literature.

### Ethics and dissemination

Formal ethical approval is not required as this study involves neither human participants nor unpublished secondary data. Findings of the scoping review will be disseminated through professional networks, conference presentations and publication in a scientific journal.

### Patient and public involvement

The design of this scoping review protocol did not involve patients. However, patients’ beliefs and experiences are central to the research question

## Data Availability

This is a protocol for a scoping review, and all data produced in the present work will be contained in the manuscript

## Authors’ contributions

JGA descent conceived the idea of this project, established the research question and methodology, and led the development of the protocol. LV and HF contributed to the methods and gave meaningful contributions to the development and/or editing of the protocol. CMEL and JGA developed the search strategies. JOA and AP helped refine the protocol. All authors have approved the final version of the protocol.

## Appendix

### 1 Medline (Ovid)

Ovid MEDLINE(R) ALL / PubMed(R) <1946 to Present>

1. “artificial intelligence”.ti,ab. or exp Artificial Intelligence/ 226908
2. Chatbot*.ti,ab. 1841
3. Musculoskeletal Diseases/ 15920
4. (bone or Orthopedic or Coxa or Osteochondritis or Hip or Knee or Ankle or Shoulder or wrist or Hand or Spine or Back or Neck or Arthritis or limb or orthop* or osteochondr* or Osteonecros* or osteoporos* or periarthriti* or polymyalgia* or scleroderma* or Spondylarthrit* or sprain or fracture* or sciatica).ti,ab. 2770180
5. 3 or 4 2778974
6. 1 or 2 227817
7. animals/ not humans/ 5218162
8. 5 and 620840
9. 8 not 7. 3565

